# Randomized controlled trial of Group Interpersonal Therapy conducted by non-specialists versus waiting list for symptoms of depression and anxiety in community young adults: a Research protocol

**DOI:** 10.1101/2025.10.13.25337768

**Authors:** Giulio Bertollo Alexandrino, Bruno Perosa Carniel, Pedro Henrico Grazziotin Portal, Graziela Nunes Peixoto, Marina Ribeiro de Matos, Luiz Carlos Nascimento da Silva, Christian Kieling, Neusa Sica da Rocha

## Abstract

**Introduction:** Depression is a highly prevalent and potentially chronic mental disorder with significant impact on quality of life and social support, with a frequent onset in early adulthood. Despite the existence of effective treatments, many individuals do not have access to care, do not respond to, or do not tolerate medications. Additionally, access to mental health services remains limited, especially in low- and middle-income countries, where the World Health Organization estimates that 75% of individuals do not receive adequate care. Group Interpersonal Therapy (IPT-G) is an evidence-based, structured psychotherapy recommended by international guidelines for the treatment of depression and is adaptable for task-shifting approaches in primary care. Previous research has shown promising results for IPT-G delivered by non-specialists in low-resource settings in an open study, but randomized trials by non-specialists are still lacking.

**Objectives:** To evaluate the improvement in depressive symptoms after eight sessions of IPT-G among young adults with depressive symptoms in a community setting, conducted by non-specialists in a randomized trial, representing a new step in demonstrating scientific evidence for novel clinical treatments.

**Methods:** This randomized controlled clinical trial is planned to include 80 participants randomized (1:1) between an interventional (IPT-G) and a control (waiting list) group. Our study will enroll young adults aged 18 to 24 years with depressive symptoms. The intervention will focus on interpersonal relationships within the IPT model and will be delivered by a university student and a medical doctor trained in IPT. The primary outcomes are depressive symptoms; secondary outcomes include quality of life and social support.

**Results:** The trial was approved by the Research Ethics Committee (CEP) of the HCPA (Project number: 2023-0283, CAAE: 73830223.7.0000.5327) and registered on ClinicalTrials.gov (NCT06480019). Recruitment started in February 2024 and is ongoing.

**Conclusion:** This study could support the integration of IPT-G delivered by non-specialists into public health systems worldwide as a low-cost and accessible intervention. Findings could help reduce mental health service gaps and increase the capacity of public health systems to treat depression.

**STRENGTHS AND LIMITATIONS OF THIS STUDY:** This study has the strength of being the first randomized controlled trial evaluating IPT-G delivered by non-specialists. It is conducted in a middle-income country, where, despite the existence of guidelines recommending IPT-G implementation by non-specialists, there are no randomized trials. The study also has some limitations: participants are not restricted from seeking other mental health treatments during the trial, and the intervention is compared with a waitlist control rather than an active treatment condition.

Version 2.0 – 10 October 2025 (Final version submitted)

## INTRODUCTION

Depression is a highly prevalent and chronic disorder with a significant impact on society. It ranked second among the leading causes of years lived with disability in 2021 (1). Global Burden of Disease (GBD) Study data (2) indicates that the estimated prevalence of depressive disorders increases significantly in the first decades of life: 0.08% in children aged 5-9 years, 0.98% in children aged 10-14 years, 2.69% in adolescents aged 15-19 years, and 3.85% in young adults aged 20-24 years. These findings highlight the critical importance of initiating preventive and therapeutic interventions during the early stages of life, when depressive disorders typically begin to manifest.

In young adults, many stressful events—such as life changes, grief, interpersonal disputes, and loneliness—can be daunting, further exacerbating their vulnerability to depression. Beyond that, depression can impair quality of life and social support, leading to academic problems, reduced work productivity, and withdrawal from social relationships (3,4). Moreover, some young adults do not exhibit all the diagnostic criteria, and many do not receive a diagnosis or appropriate treatment, or they avoid seeking treatment because of stigma and discrimination (5). The WHO estimates that 75% of people in low- and middle-income countries do not have access to treatment. To address this gap, WHO created the Mental Health Gap Action Program (mhGAP), which proposes psychological treatments for depression such as Interpersonal Therapy (IPT) and Cognitive-Behavioral Therapy (CBT) (6).

IPT is an intervention that has demonstrated to be trainable at scale and has demonstrated positive results in the treatment of depression and other conditions; however it requires trained personnel to deliver the intervention (7). The main idea of this therapy is the connection between an individual’s mood and interpersonal relations. Stressful life events (grief, interpersonal disputes, life changes, loneliness/social isolation) can be triggers for depression, and IPT works on these difficulties to help people cope with these events.

Because of the high prevalence of mental health conditions and limited access to mental health providers, WHO adopted a ‘task-shifting’ or ‘task-sharing’ approach, in which traditionally specialized health care tasks are delegated to and/or shared with existing or new staff with less training or training narrowly tailored to the service required. These approaches involve transferring basic responsibilities to non-specialists to reduce the population burden and free up specialized health resources for more complex tasks (8).

Mutamba et al. in Uganda—a country where the public health system allocates very scarce resources to mental health—conducted a non-randomized clinical trial in which caregivers from certain villages received the usual psychotherapy treatment, whereas another group of caregivers received, in addition, 12 sessions of group IPT (IPT-G) conducted by lay community workers in the context of primary care. The objective was to evaluate the effectiveness of this therapy in reducing depression among caregivers of children with Nodding Syndrome, a neuropsychiatric condition with high morbidity and mortality. Caregivers in IPT-G, and their children, had a significantly greater reduction in depressive symptoms compared with caregivers who received only the usual treatment (9). These results strengthen the evidence that mental health interventions in primary care, even when conducted by non-specialized professionals, can bring numerous health benefits to the population.

To establish evidence-based psychotherapy approaches, some steps are required: theoretical development, pilot studies, randomized controlled trials, systematic reviews and meta-analyses, replication in diverse contexts, and clinical guidelines integration (10). Although the WHO already has an IPT-G manual for training non-specialists, randomized clinical trials (RCTs) are needed to demonstrate scientific evidence and support the implementation of this therapy worldwide. Thus, this study aims to evaluate the effectiveness of IPT applied by non-specialists in reducing depressive symptoms compared with a waiting list in young people in a pragmatic RCT, aiming to investigate the potential of this approach as a powerful ally in the prevention and treatment of this disorder (11). The main outcomes are depressive and anxiety symptoms, and the secondary outcomes are quality of life, social support, therapy adherence, and perception of clinical improvement. It is hypothesized that young adults who receive therapy from non-specialists will show improvements in the depressive and anxiety symptoms, social support, and quality of life compared with participants on the waiting list.

## METHODS

### Design

The trial is designed as a pragmatic, controlled, randomized, and longitudinal study, comprising an intervention group (experimental) and a control group (waiting list). The intervention group will receive an IPT-G intervention delivered by non-specialists, following an 8-week protocol.

### Randomization

Participants will be randomly allocated to the intervention or control group in a 1:1 ratio using simple randomization generated by the Research Electronic Data Capture (REDCap) platform (12,13). Allocation is computer-generated by an independent researcher who is not involved in participant recruitment, enrollment, or intervention delivery. After every 20 new participants complete baseline assessments, randomization determines who will be assigned to the intervention or to the wait-list control group. The intervention team (therapists) and data collection team operate independently, with no interaction between them. Therapists are blinded from the participants’ assessments data to minimize bias and data collectors will only have access to the instruments once therapy is complete. Given the nature of the intervention, participants are not blinded to allocation, and the therapists have no access to information about the control group.

### Ethical procedures

This study was approved by the Research Ethics Committee of Hospital de Clínicas de Porto Alegre (Project number: 2023-0283, CAAE: 73830223.7.0000.5327). The eligible population will be invited to participate in the study without any constraint and, upon voluntary acceptance, will sign an informed consent form, in accordance with Resolution no. 466 of the National Health Council. Any risk situations (suicidal ideation or risk, psychotic symptoms, or mania) will result in caregivers being contacted, and the participant referred to emergency medical services.

### Study Setting

This project will be conducted in the city of Porto Alegre, in the state of Rio Grande do Sul. The meetings will be held at the Luis Guedes Study Center (CELG), an establishment linked to the Hospital de Clínicas de Porto Alegre (HCPA). The enrolled population comprises a very heterogeneous group, reflecting the city’s diverse socioeconomic scenarios. According to Vigitel 2021, Porto Alegre had the highest prevalence of depression among all Brazilian capitals (17.5%), compared with the national average of 11.3% (14).

Our team consists of the principal researcher, psychiatrists, psychologists, medical doctors, and medical students. Medical doctors and students received IPT training from the principal researcher for 3 months, followed by the test to certify the knowledge acquired. The test questions were obtained from the Group Interpersonal Therapy (IPT-G) Manual developed by the World Health Organization (11). The questions are available in the annex of the WHO manual and were also included as an appendix 1 in the present article.

Data collection begins at inclusion, continues during the 8-week intervention, and is followed up after 6 months. For the primary outcome, all patients are followed until six months after the last therapy session. Recruitment began in March 2024 and will continue until the end of 2025. Evaluations take place at baseline (initial interview), after each session, and during follow-up. Participants are invited via social media, journals, e-mail, and posters.

### Patient and Public Involvement

Patients, researchers, and specialists are involved at various stages of the trial, including its design, management, and conduct. Our study is a community-based clinical trial, conducted in collaboration with the intervention team and research team, who work independently. Some strategies were incorporated into our study:

a. Design and conduct: Participants’ feedback is being considered in the planning of group sessions and the structure of the intervention. The supervision of therapy occurs on a weekly basis, and participants’ interests are continually considered during the intervention.
b. Choice of outcome measures: Outcome measures were selected based on their relevance to participants’ mental health experiences and the type of therapy (depressive and anxiety symptoms, social support, and quality of life).
c. Recruitment: Recruitment strategies were designed to recruit young adults with depression, utilizing communication channels and language appropriate to the target population. An email is available to ask questions about the project.

Our main results will be discussed with all stakeholders (patients, researchers, and specialists) involved in the project, and we will seek to develop together the most appropriate method of dissemination to the broader community.

### Recruitment setting and population

Young adults aged 18–24 years who live or work in Porto Alegre (Brazil) are being recruited. Recruitment occurs via social media (Instagram, Facebook, WhatsApp) and printed posters placed at Primary Care Units, universities, and preparatory-course venues within the metropolitan area.

Individuals interested in participating will be invited to access REDCap (12,13) link and meet the pre-screening criteria:

a. Live, work, or study in the geographic area of Porto Alegre
b. Express a desire to voluntarily participate in the research and sign an informed consent form
c. Be aged 18 to 24 years.
d. Be able to attend all 8 meetings.
e. Score ≥5 on the Patient Health Questionnaire (PHQ-9), without indication of suicidal ideation as assessed by item 9 (≤ 1) of the scale.

### Pre-screening and safety

The online form includes the PHQ-9 to identify depressive symptomatology (PHQ-9 ≥ 5). Suicidality is assessed using PHQ-9 item 9. Respondents endorsing suicidal ideation (any response ≥2) will not proceed and will immediately receive a standardized message with emergency contacts and local referral options within the online recruitment platform.

### Eligibility assessment

Candidates who pass pre-screening will be invited (by phone, WhatsApp, or email) to attend a structured clinical interview to confirm inclusion/exclusion criteria. Screening outcomes (eligible/ineligible and reasons) will be being recorded.

A trained medical doctor will apply the socio demographic questions and the Brazilian Portuguese version of the Mini International Neuropsychiatric Interview (MINI)(15). Individuals presenting with the following conditions will be excluded, as diagnosed by the MINI following the DSM-IV criteria:

a. Manic or hypomanic episode (current)
b. Psychotic syndrome (current or past)
c. Substance dependence or substance abuse (last 12 months, except tobacco/nicotine)
d. Moderate or high suicide risk, as operationalized by the MINI
e. Diagnosis of other types of depression, such as bipolar disorder

If an individual shows suicidal risk during in-person enrollment or the clinical trial, they will be referred to emergency care. These individuals will be excluded from the trial.

### Participant Timeline

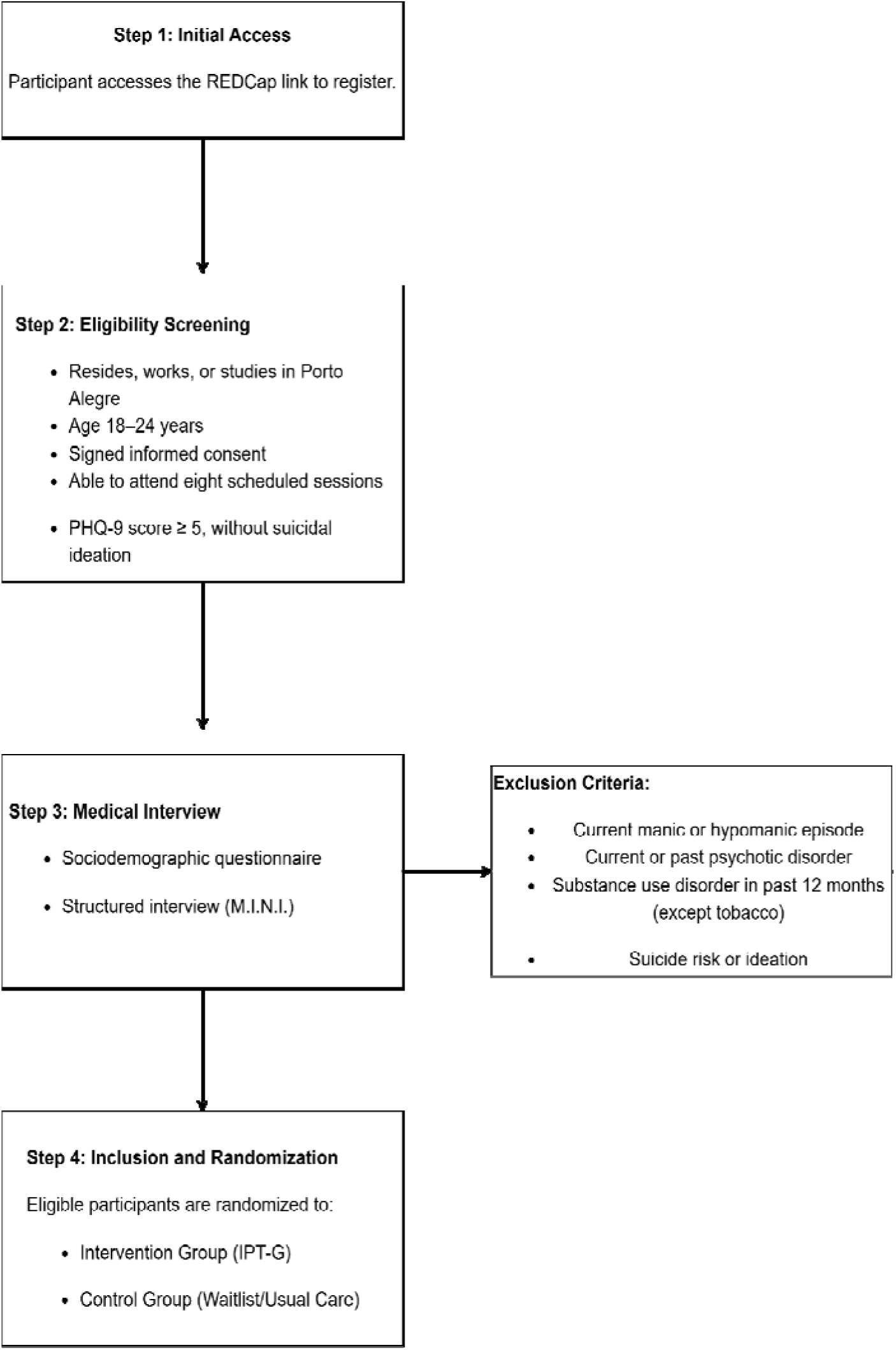

### Identification and approach

Interested individuals access an ethics-approved advertisement containing a link to an online pre-screening form. Research staff (psychiatrists, psychologists, medical doctors, and medical students) monitor responses and contact potential participants who appear eligible. Recruitment materials have been approved by the Ethics Committee.

### Informed consent

Eligible participants receive an informed consent form and have adequate time to ask questions. Consent is obtained prior to any baseline procedures. Participation is voluntary and may be discontinued at any time. Any important protocol modifications will be submitted to the Research Ethics Committee for review and approval before implementation. Participants will be informed of any changes that may affect their participation, and re-consent will be obtained when required by the Committee.

### Compensation and costs

Participation involves no costs to participants, and no financial compensation is offered.

### Enrollment and allocation

After obtaining consent, completing socio-demographic and MINI interviews and completing baseline measures, participants who meet all criteria are enrolled and scheduled for randomization.

### Intervention

The intervention was proposed based on IPT and will focus on one problem area: grief; interpersonal disputes; role transitions; or interpersonal deficits (7,11). The 4 IPT areas are:

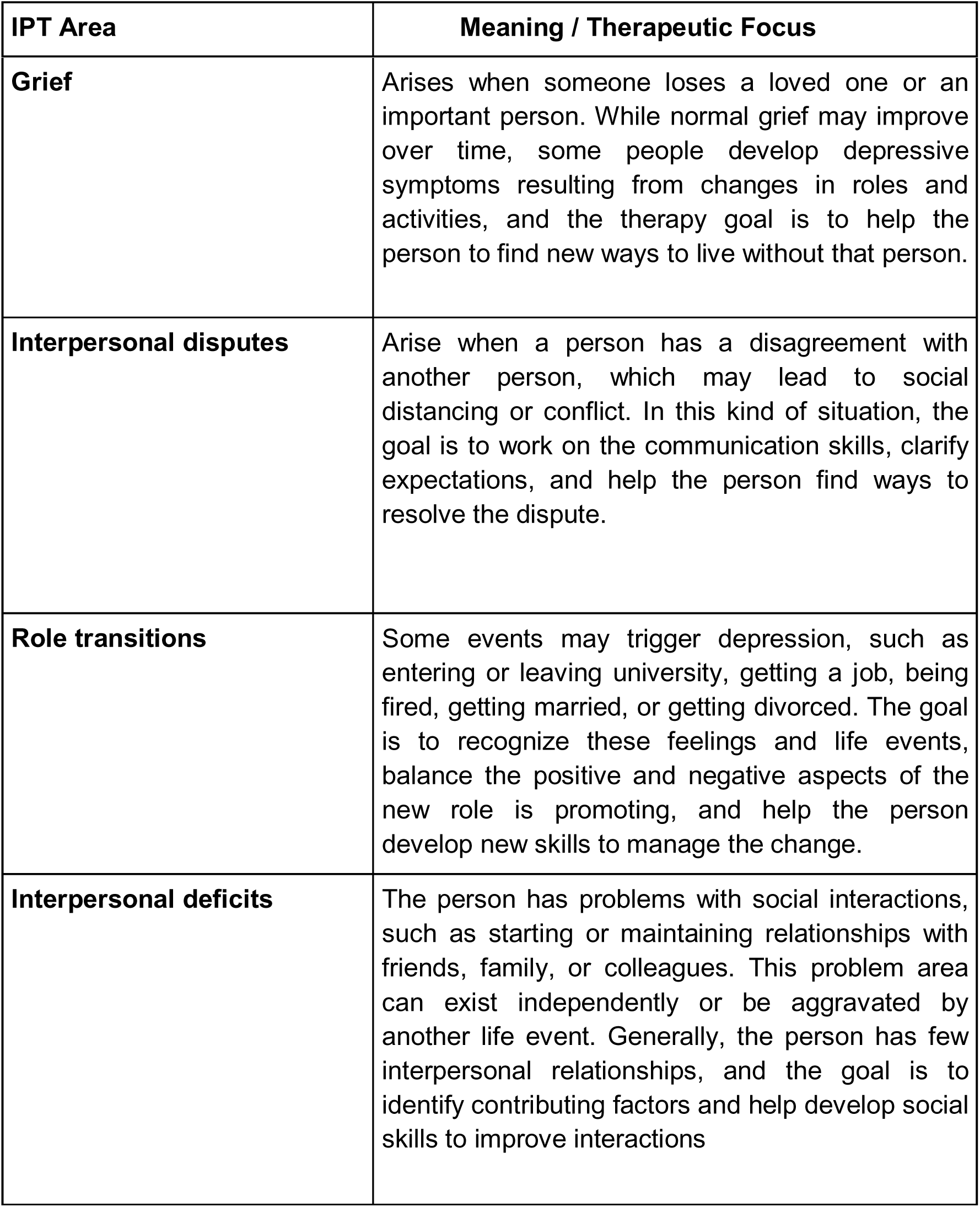

It will consist of 8 weekly meetings—one individual and seven group sessions. The individual meeting will last one hour, and the group sessions about 2 hours. Each group will have at least 5 and no more than 10 participants. The therapists will be a medical student and a medical doctor without prior specialization in IPT. The principal investigator will be responsible for training and supervising the medical students and doctors during all group sessions. All sessions will be audio-recorded so they can be evaluated later, ensuring that the meeting mediators have covered all the main points of problem-solving therapy.

An initial session (the individual session) will be held to assess the occurrence of depressive symptoms through anamnesis. An interpersonal inventory will also be conducted to evaluate how current interpersonal relationships may be related to the participant’s depressive symptoms. The identification of satisfactory and unsatisfactory aspects of life will also be included, as well as clarification of what IPT is, establishment of a therapy contract, and identification of each participant’s IPT problem area.

In the intermediate sessions (2nd to 7th), the problem areas are addressed, such as grief, interpersonal disputes, role transition, and interpersonal deficits. At this stage, strategies are applied to achieve the objectives defined for the problem areas. Although more than one area may be present, the intervention will focus on the one most relevant to current depressive symptoms.

The last session will consist of a discussion about the end of the intervention, recognition that the end of the intervention is a grieving process, recognition of the competence and independence of the patient/participant, review of gains from the intervention, and planning for continuation and maintenance. The possibility of lack of response to the proposed intervention will also be addressed, with referral to other interventions.

| Session | Key Points |
| --- | --- |
| Initial Session (1st) | • Anamnesis and assessment of depressive symptoms<br>• Introduction to IPT<br>• Interpersonal inventory<br>• Identification of problem area<br>• Therapy contract |
| Intermediate Sessions (2nd-7th) | • Address problem areas:<br>- Grief<br>- Interpersonal disputes<br>- Role transitions<br>- Interpersonal deficits<br>• Apply specific strategies for the most relevant area |
| Final Session (8th) | • Discussion of therapy end as a grieving process<br>• Recognize competence and independence<br>• Highlight gains and plan for future maintenance<br>• Address lack of response to intervention |

### Follow-up Session

All participants in the intervention group will be invited to a meeting at HCPA six months after therapy to discuss what happened after the sessions, specifically how they implemented the strategies in their lives and how they are feeling about their life goals. At this stage, the study will remain blinded to group allocation for outcome assessment purposes.

### Confidentiality and data handling

Contact details are stored separately from research data in a secure, access-controlled REDCap system. Only authorized study staff have access. Communications via WhatsApp follow the institution’s privacy procedures. Aggregate screening numbers (assessed, excluded with reasons, enrolled) are reported.

### Retention strategies

To minimize participant loss, appointment reminders are sent (SMS/WhatsApp/email). Flexible scheduling is offered during enrollment. Participants who miss visits are contacted by telephone or WhatsApp, when deemed necessary by the supervisor. Moreover, a WhatsApp group is created to send reminders when all group members are involved.

### Ancillary and post-trial care

A physician will be present in all sessions. If a participant shows suicidal risk, the physician will contact a family member to come to the hospital, and the participant will be referred to psychiatric emergency services. After the intervention group completes therapy, the waiting-list control group will be offered IPT-G if results are favorable. If a participant shows worsening symptoms, our medical team and psychiatrists will provide immediate treatment and refer them to the public health system.

### Alternative care and referrals

Participants are allowed to receive concomitant care. Individuals who decline or withdraw are provided with information on public mental health services and community resources for support and treatment.

### Harms

Given that the intervention consists of IPT-G delivered by trained non-specialists, no pharmacological or invasive procedures are involved. Therefore, the expected risk to participants is minimal.

### Outcomes

The primary outcome of the study will be depressive and anxiety symptoms, as these represent the main targets of IPT-G and are the most clinically relevant indicators of improvement in young adults with depressive symptomatology.

The secondary outcomes of this study will be quality of life and perceived social support. Depressive symptoms frequently impair functioning and perceived quality of life (QoL). In addition, IPT-G emphasizes interpersonal relations and enhancement of social support, which may act as a key mediator of the intervention effects. To investigate moderators, mediators, and potential adverse effects, the following constructs will be measured: general health status, resilience, severity of the depressive episode, adverse effects of psychotherapy, attachment style, personality traits, and history of childhood trauma.

### Data Collection

We will evaluate these outcomes at different moments.

- Baseline (post-inclusion): All participants will complete the full set of questionnaires before randomization.
- During the intervention: At the end of each IPT-G session, all participants will complete the questionnaires to capture short-term fluctuations in mood and adverse events.
- Post-intervention and follow-up: Full assessments will be administered immediately after the last group session and at the 6-month follow-up.

This structure enables both longitudinal analysis of the intervention and exploratory modeling of how associated factors influence treatment effects. Table 1 presents the structure of our data collection.

**Table 1.** Data Collection Structure.

| Timepoint | Enrollment | Allocation | Post-allocation (sessions 1–6) | Post-intervention (8 weeks) | Follow-up (6 months) |
| --- | --- | --- | --- | --- | --- |
| Enrollment |  |  |  |  |  |
| Eligibility screen | X |  |  |  |  |
| Informed consent | X |  |  |  |  |
| Baseline measures | X |  |  |  |  |
| Randomization |  |  |  |  |  |
| Intervention: IPT-G |  | X |  |  |  |
| Control: Waiting list |  | X |  |  |  |
| Assessments |  |  |  |  |  |
| HADS, PHQ-9, BDI | X |  | X | X | X |
| WHOQOL-BREF | X |  | X | X | X |
| MOS-SSS | X |  | X | X | X |
| CGI |  |  | X | X | X |
| CD-RISC-10 | X |  | X | X | X |
| RSQ | X |  |  | X |  |
| CTQ | X |  |  | X |  |
| EPQR-A | X |  | X | X | X |
| GHQ-12 | X |  | X | X | X |
| UE-ATR |  |  | X | X | X |

### Instruments

#### Screening

The PHQ-9 is a self-administered instrument that quantitatively assesses the severity of depressive symptoms over the past two weeks (16). It consists of nine items corresponding to the diagnostic criteria for major depressive disorder in the DSM-IV. Each item is scored on a 4-point Likert scale (0 = not at all to 3 = nearly every day), resulting in a score ranging from 0 to 27. In a Brazilian sample, the version translated into Brazilian Portuguese demonstrated validity of its psychometric properties (17). A score greater than or equal to 5 was used as the inclusion criteria.

### Primary Outcomes

#### Depression and Anxiety

The Hospital Anxiety and Depression Scale (HADS) consists of 14 items, of which 7 assess anxiety and 7 assess depression. Each item is scored from 0 to 3, and each subscale has a maximum score of 21 points with higher scores indicating greater symptom severity (18). In a Brazilian sample, the version translated into Brazilian Portuguese demonstrated validity of its psychometric properties (19).

### Secondary Outcomes

#### 1- Quality of Life

The World Health Organization Quality of Life—BREF (WHOQOL-BREF) is a scale with 26 items that measure quality of life. It includes two general items and 24 items representing four domains (physical, psychological, social relationships, and environmental) (20). In a Brazilian sample, the version translated into Brazilian Portuguese demonstrated validity of its psychometric properties (21).

#### 2- Social Support

The Medical Outcomes Study Social Support Survey (MOS-SSS) measures each individual’s perceptions of personal support. The scale comprises five support factors (emotional, informational, material, affective, and positive social interaction). Higher scores indicate greater perceived support (22). In a Brazilian sample, the version translated into Brazilian Portuguese demonstrated validity of its psychometric properties (23).

#### 3- Depression Symptoms

Secondary outcomes include depressive symptoms assessed by the PHQ-9 (also used for screening) and the Beck Depression Inventory (BDI), which measure the same construct as the primary outcome (depressive symptoms). Both instruments were incorporated in our trial as complementary scales to allow a more comprehensive analysis of depressive symptomatology.

The PHQ-9 is a self-administered instrument that quantitatively assesses the severity of depressive symptoms over the past two weeks(16). It consists of nine items corresponding to the diagnostic criteria for major depressive disorder in the DSM-IV. Each item is scored on a 4-point Likert scale (0 = not at all to 3 = nearly every day), resulting in a score ranging from 0 to 27. In a Brazilian sample, the version translated into Brazilian Portuguese demonstrated validity of its psychometric properties (17). A score greater than or equal to 5 was used as the inclusion criteria.

The BDI is a self-report instrument and assesses the presence and severity of depressive symptoms. It consists of 21 items describing cognitive, affective, and somatic manifestations of depression. Each item is scored in a Likert scale ranging from 0 to 3, varying from 0 to 63 points (24). In a Brazilian sample, the version translated into Brazilian Portuguese demonstrated validity of its psychometric properties (25).

#### 4- Clinical Impressions

The Clinical Global Impressions (CGI) scale is an instrument that enables clinicians to assess the overall severity of the illness, the change in clinical status over time, and the effectiveness of the intervention used. Its score can range from 1 (not ill) to 7 (extremely ill)(26). In a Brazilian sample, the version translated into Brazilian Portuguese demonstrated validity of its psychometric properties (27).

### Moderating variables

#### 1- Attachment

The Relationship Scales Questionnaire (RSQ) assesses attachment styles in interpersonal relationships. It consists of 30 items describing thoughts and feelings in close relationships. Respondents rate their agreement with each item on a 5-point Likert scale(28). In a Brazilian sample, the version translated into Brazilian Portuguese demonstrated validity of its psychometric properties(29).

#### 2- Childhood Trauma

The Childhood Trauma Questionnaire (CTQ) is a retrospective self-report instrument designed to assess experiences of childhood abuse and neglect. It consists of 28 items scored on a 5-point Likert scale, measuring five domains of maltreatment: emotional abuse, physical abuse, sexual abuse, emotional neglect, and physical neglect(30). In a Brazilian sample, the version translated into Brazilian Portuguese demonstrated validity of its psychometric properties (31,32).

#### 3- Resilience

The 10-item Connor-Davidson Resilience Scale (CD-RISC-10) is an abbreviated version of the original 25-item scale. It measures the capacity to adapt physically, mentally, and spiritually to the circumstances imposed by life (33). The 10 items are scored from 0 to 4, yielding a total score from 0 to 40. Higher scores indicate higher levels of resilience. In a Brazilian sample, the version translated into Brazilian Portuguese demonstrated validity of its psychometric properties (34).

#### 4- Personality

The Eysenck Personality Questionnaire–Revised (EPQR-A) is a self-administered questionnaire composed of 24 items divided into four domains: the three dimensions of personality (neuroticism, extraversion, and psychoticism) and a validity scale (social desirability). This instrument is a shorter and updated version of the original 90-item scale(35). In a Brazilian sample, the version translated into Brazilian Portuguese demonstrated validity of its psychometric properties (36).

### Associated Factors

#### 1- Adverse Effects of Psychotherapy

The UE-ATR checklist is an instrument that enables finding, classifying, and evaluating adverse effects of psychotherapy, discriminating between undesirable effects, adverse reactions to treatment and side effects. It lists 17 domains of possible undesirable effects, which are evaluated on a 1 to 5 severity scale and for their relationship with psychotherapeutic treatment (37). Although there is a translation into Brazilian Portuguese, its psychometric validation in Brazilian samples has not yet been published.

### Sample Size

A total sample size of 68 participants (34 per group) was estimated to detect a minimum difference of 3 points in the mean HADS scores between the Intervention and Control groups at 8 weeks. This threshold was chosen because a difference of ≥3 points on the HADS has been considered clinically meaningful in previous research on depression and anxiety interventions. Allowing for an additional 10% to account for potential attrition and refusals, the final required sample size is 76 participants. The estimation assumed a statistical power of 80%, a two-sided significance level of 5%, and a standard deviation of 4 points(38). Expected retention rates were 90% at time point 2 and 80% at time point 3. An autoregressive correlation matrix with parameter 0.5 was incorporated into the model. The sample size calculation was performed using the PSS Health online tool(39).

### Statistical Analysis

First, the data will be reviewed for missing information. Our study will use a linear mixed-effects model (LMM) for the study’s main outcomes, such as depressive and anxious symptoms, quality of life, and social support with fixed effects for Group, Time, and Group x Time, adjustment for baseline values of each outcome and relevant clinical/demographic covariates, and a participant random intercept. This analysis type was chosen due to potential study limitations, such as loss of participants during therapy in both groups and missing data. Clinical trials tend to present losses, and psychotherapy research often shows dropouts during treatment, leading to loss of data and power. The LMM framework enables the inclusion of all available observations under a missing-at-random (MAR) assumption and avoids ad-hoc imputations (e.g., LOCF), which can bias estimates and reduce power(40). For our primary outcomes, sensitivity analyses will be performed using multiple imputation, median imputation sensitivity, and per-protocol analysis (attendance ≥4 sessions and post-intervention assessment); these results will be contrasted with the LMM. Clinical and demographic characteristics will be included as explanatory variables. All data will be analyzed using SPSS v.30 software(41).

### Data Monitoring

A formal independent Data Monitoring Committee was not established for this trial. The intervention is a low-risk, non-pharmacological psychological treatment, and does not involve medical, invasive, or pharmacological procedures that would typically require independent data and safety monitoring. Trial conduct is monitored weekly by the principal and co-investigator. The study will proceed until completion unless ethical or safety concerns arise, in which case the principal investigator and the research steering committee will decide on suspension or termination in consultation with the Research Ethics Committee.

## TRIAL STATUS

Our trial was registered at ClinicalTrials.gov (NCT06480019 - https://clinicaltrials.gov/study/NCT06480019). Recruitment started in February 2024 and, at the time of submission of this protocol’s manuscript, recruitment is ongoing, and the first groups have already commenced.

## DISCUSSION

Previous studies have demonstrated the efficacy of IPT in treating depressive symptoms when conducted by specialized professionals (7,42). However, some patients are medication non-responders and lack access to psychotherapy (43). In public health settings, individual psychotherapy services are also scarce. Furthermore, the most prevalent psychotherapy modalities, such as analytic and cognitive behavioral therapies, require years of training and typically involve multiple treatment sessions (44). In contrast, IPT is relatively straightforward to learn, auto limited, and has been shown to be effective for depressive symptoms when conducted by non-specialists in open clinical trials (45). Our study aims to substantiate its effectiveness in a randomized trial, representing the next step in generating scientific evidence for novel clinical treatments. Should the study yield positive results, IPT could be effectively implemented in public health systems globally, reducing mental health gap, lowering costs, increasing treatment capacity, and improving access for the population.

## Supporting information

SPIRIT Guideline

## Data Availability

All data supporting the findings of this study protocol are included in this article and its supplementary materials. Other data will be available from the authors upon reasonable request in order to protect participant s anonymity and privacy.

## Dissemination policy

After data analysis, all participants and community stakeholders will be invited to a results presentation. No identifiable information will be disclosed. Findings will be disseminated through journal articles and presentations at national and international scientific conferences. Preprints may be used to accelerate access, with subsequent journal publication following peer review. Reporting will follow CONSORT guidelines.

## Consent for publication

All participants involved in this study signed an informed consent form, explicitly authorizing the collection, use, and publication of their data for scientific and academic purposes.

## Availability of data and materials

All data supporting the findings of this study protocol are included in this article and its supplementary materials. Other data will be available from the authors upon reasonable request in order to protect participant’s anonymity and privacy.

## Author Contributions

NR and CK conceptualized the project. NR developed and coordinated the project at HCPA and was responsible for grant acquisition. GA, BC, and PD are responsible for implementing and conducting the project. BC and PD are medical doctors; GP, MM and PP are medical students who conducted the therapy. GA and LS collected data. Part of this project will be the subject of GA’s PhD dissertation.

## Acknowledgements

We thank the Centro de Estudos Luís Guedes and the Department of Psychiatry at the Hospital de Clínicas de Porto Alegre (HCPA) for providing facilities to conduct the therapy sessions. We also thank all professionals, collaborators, and participants who made this study possible.

## Sponsor

Hospital de Clínicas de Porto Alegre (HCPA).

Rua Ramiro Barcelos, 2350 – Porto Alegre, RS, Brazil – CEP 90035-903

Phone: +55 (51) 3359-8000

Website: www.hcpa.edu.br

Conselho Nacional de Desenvolvimento Científico e Técnológico (CNPq).

Setor de Autarquias Sul (SAUS), Quadra 1, Lotes 1 e 6, Edifício Telemundi II, Bloco H – Brasília, DF – CEP 70070-010

Phone: +55 61 3211-4000

Website: https://www.gov.br/cnpq/pt-br

Coordenação de Aperfeiçoamento de Pessoal de Nível Superior – Brasil (CAPES).

Setor Bancário Norte, bloco L, lote 6, 4° andar - Brasília, DF - CEP 70040-020

Phone: +55 800 616161

Website: https://www.gov.br/capes/pt-br

## Appendix 1

### Group IPT Knowledge Test

1. Please name the four types of triggers of depression in IPT define each one.
2. What are the phases of Group IPT? (Just mention them.)
3. Describe the tasks that need to take place in the pre-group phase.
4. Describe the goals and steps of the initial group phase.
5. Describe how a typical session starts in the middle phase.
6. In the middle phase, what are the strategies for working with grief?
7. What are the stages of a dispute? (Please describe each.)
8. What are the strategies for managing disputes?
9. What are the strategies for life changes?
10. What are the strategies for loneliness/social isolation?
11. What is communication analysis? Can you give an example?
12. Describe 2 other techniques that you use in Group IPT (across strategies). Please explain.
13. Describe 1 way to involve group members in group discussions, and give examples.
14. What is something you can do to distribute your time fairly among the group members?
15. What happens during the termination phase?

Test scoring instructions are as follows: every question gets 2 points if answered adequately, 1 point if something important is missing and 0 points if the answer is incorrect. Usually the facilitator should be able to answer 70% of the 15 questions correctly (21 points).

### Participant Timeline

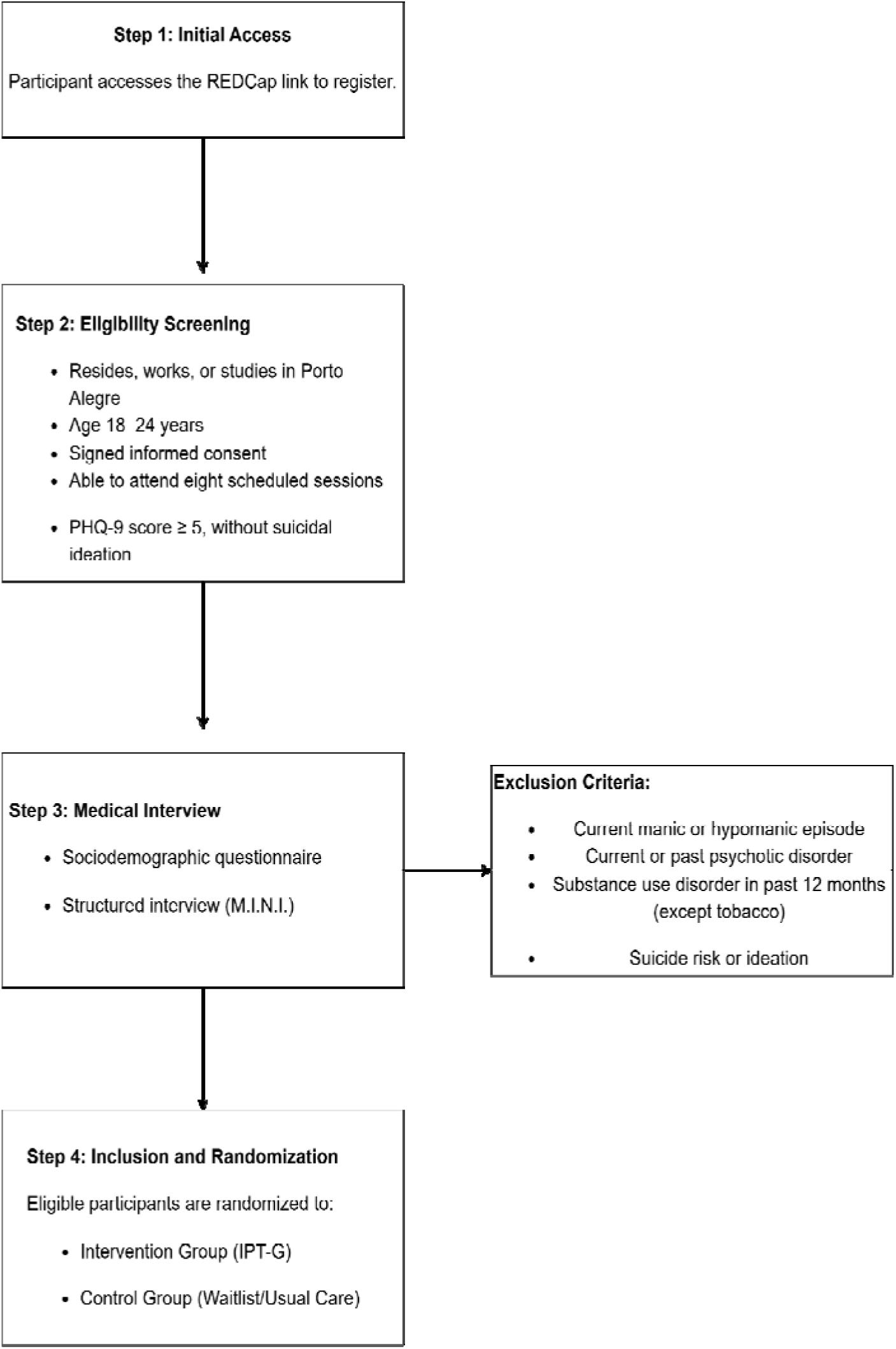

